# Gut microbiome dysbiosis and immune activation correlate with somatic and neuropsychiatric symptoms in COVID-19 patients

**DOI:** 10.1101/2024.11.18.24317428

**Authors:** Paula L. Scalzo, Austin Marshall, Sirena Soriano, Kristen Curry, Mario Dulay, Timea Hodics, Eamonn MM Quigley, Todd J. Treangen, María M. Piskorz, Sonia Villapol

## Abstract

COVID-19 patients often exhibit altered immune responses and neuropsychiatric symptoms during hospitalization. However, the potential interactions with gut microbiome profiles have not been fully characterized. Here, COVID-19 disease severity was classified as low (27.4%), moderate (29.8%), and critical (42.8%). Fever (66.1%) and cough (55.6%) were common symptoms. Additionally, 27.3% reported somatic symptoms, 27.3% experienced anxiety, 39% had depressive symptoms, and 80.5% reported stress. Gut microbiome profiling was performed using full-length 16S rRNA gene sequencing. Elevated interleukin-6 levels were observed in the most severe cases, indicating systemic inflammation. Reduced gut bacterial diversity was more pronounced in women and obese patients and correlated with higher disease severity. The presence of the genus *Mitsuokella* was significantly associated with increased physical, stress, anxiety, and depressive symptoms, and *Granulicatella* with critically ill patients. These findings suggest a link between mental health status, systemic inflammation, and gut dysbiosis in COVID-19 patients, emphasizing the potential of microbiome-targeted therapies to improve recovery and reduce severe complications.

## Introduction

Coronavirus disease 2019 (COVID-19), caused by the severe acute respiratory syndrome coronavirus 2 (SARS-CoV-2), primarily presents with respiratory symptoms. However, the infection can also affect neurological(*1*) and gastrointestinal (GI) systems(*2, 3*). It is well established that men are at a higher risk than women of developing severe acute COVID-19(*4, 5*), as well as individuals over the age of 60 years old(*6, 7*). Patients hospitalized during the acute phase of COVID-19 exhibit a high prevalence of mental health issues such as elevated stress, anxiety, and depression(*8–10*). These mental health disturbances are more prevalent in women and can be worsened by factors such as age(*11*), more severe physical symptoms, or length of hospital stay(*12*). Furthermore, the presence of COVID-19 symptoms at the time of admission adds to the psychological burden(*13*).

SARS-CoV-2 infection increases levels of soluble immune mediators in the bloodstream, such as inflammation-related cytokines(*14*). It activates intestinal angiotensin-converting enzyme 2 (ACE2) receptors(*15*) and damages the intestinal epithelium, disrupting the gut barrier(*16, 17*), as observed in patients with severe COVID-19(*18, 19*). This inflammatory response triggers GI and alterations in the gut microbiome(*19–21*), both of which are associated with disease severity.

Gut dysbiosis, characterized by a reduction in butyrate-producing, anti-inflammatory bacteria, and increased pro-inflammatory taxa, disrupts immune regulation, nutrient metabolism, and structural defenses, contributes to systemic inflammation and impairs host homeostasis(*22*). Microbiome imbalances during the acute phase of COVID-19, especially in hospitalized patients, were linked to increased mortality rates(*23*). Previous studies have identified microbial features in the gut and airways of COVID-19 patients during hospitalization and recovery(*21*), suggesting that microbial markers could serve as noninvasive diagnostic tools. Microbiome dysbiosis also affects immune and inflammatory response regulation and brain function(*24*). However, the link between microbial alterations and somatic or neuropsychiatric symptoms in COVID-19 patients and their potential as predictive tools has yet to be fully elucidated.

In this cross-sectional study, we identified associations between somatic and neuropsychiatric symptoms, inflammatory profiles, and alterations in gut microbiota composition in hospitalized COVID-19 patients. By examining these relationships, we aim to gain insights into the mechanisms underlying COVID-19 pathogenesis and potentially identify novel therapeutic targets for intervention.

## Results

### Characteristics of the participants

We enrolled 124 COVID-19 patients, 63 males and 61 females. The mean age was 55.2 (±14.6) years. The average length of hospital stay was 7 days (range, 1 to 21 days), and a total of 8 (6.5%) patients died during their initial hospital stay. According to the COVID-19 Severity Index, 34 (27.4%) patients were classified as low, 37 (29.8%) as moderate, and 53 (42.8%) as critical COVID-19. Older patients experienced more severe COVID-19 symptoms (p < 0.001), and the prevalence of men increased with the severity of the disease (p = 0.025). Data are summarized in **Table 1**.

**Table 1.** Demographic and clinical characteristics of the patients classified according to the COVID-19 Severity Index (n=124).

| Variables | All patients<br>(n=124) | COVID-19 Severity |  |  | p Value |
| --- | --- | --- | --- | --- | --- |
|  |  | Low<br>(n=34) | Moderate<br>(n=37) | Critical<br>(n=53) |  |
| Age, years, mean ± SD | 55.2 ± 14.6 | 42.3 ± 11.1 | 55.5 ± 12.8 | 63.2 ± 11.6 | <0.001<sup>#</sup> |
| <60 years, n (%) | 78 (62.9) | 34 (100) | 24 (64.9) | 20 (37.7) |  |
| ≥60 years, n (%) | 46 (37.1) | 0 | 13 (35.1) | 33 (62.3) | |
| Gender, n (%) | | | | | 0.025<sup>\$</sup> |
| Male | 63 (50.8) | 12 (35.3) | 17 (45.9) | 34 (64.2) |  |
| Female | 61 (49.2) | 22 (64.7) | 20 (54.1) | 19 (35.8) |  |
| Body Mass Index (BMI), kg/m<sup>2</sup>, mean ± SD | 30.7 ± 6.6 | 28.7 ± 4.7 | 30.5 ± 6.2 | 32.2 ± 7.5 | 0.050<sup>#</sup> |
| <30 kg/m<sup>2</sup> | 67 (54.0) | 24 (70.6) | 20 (54.1) | 23 (43.4) |  |
| ≥30 kg/m<sup>2</sup> | 57 (46.0) | 10 (29.4) | 17 (45.9) | 30 (56.6) | |
| Co-morbidities, n (%) |  |  |  |  |  |
| Diabetes mellitus | 4 (3.2) | 0 | 2 (5.4) | 2 (3.8) | 0.570* |
| Heart diseases | 5 (4.0) | 0 | 0 | 5 (9.4) | 0.044* |
| Hypertension | 38 (30.6) | 5 (14.7) | 12 (32.4) | 21 (39.6) | 0.047<sup>\$</sup> |
| Chronic pulmonary obstructive disease | 8 (6.5) | 0 | 1 (2.7) | 7 (13.2) | 0.032* |
| Days of hospitalization, median (IQR) | 7 (4 – 9) | 7 (4 – 8) | 6 (4 – 8.5) | 7 (4 – 10.5) | 0.455<sup>&</sup> |
| Intensive care unit (ICU) | 13 (10.5) | 0 | 2 (5.4) | 11 (20.8) | <0.001* |
| Supplemental oxygen, n (%) | 58 (46.8) | 0 | 8 (21.6) | 50 (94.3) | <0.001<sup>\$</sup> |
| Antibiotics, n (%) | 25 (20.2) | 3 (8.8) | 9 (24.3) | 13 (24.5) | 0.154<sup>\$</sup> |
| Death during hospitalization, n (%) | 8 (6.5) | 0 | 1 (2.7) | 7 (13.2) | 0.032* |
| Number of COVID-19 symptoms | 3 (2 – 4) | 3 (2 – 4) | 3 (2 – 3) | 3 (2 – 4) | 0.287<sup>&</sup> |

**Note:** Data were presented using absolute values and percentages (%), mean and standard deviation (SD) or median and interquartile range (IQR). <sup>\$</sup>Chi square test. \*Fisher's exact test (FET). <sup>#</sup>One-way ANOVA. <sup>&</sup>Kruskal-Wallis test.

### COVID-19 symptoms, somatic and neuropsychiatric assessments, and inflammatory profile

Patients (n=43, 34.7%) experienced at least 3 of the 16 assessed COVID-19 symptoms at baseline. Fever (n=82, 66.1%) and cough (n=69, 55.6%) were the most common symptoms. A significant association was found between COVID-19 severity and the presence of dyspnea (p<0.001). The distribution of symptoms at the time of hospital admission, categorized by COVID-19 severity, is represented in **Fig. 1a**. Among the 77 patients who completed the instruments, 21 (27.3%) were classified as having severe somatic symptoms as intense and persistent pain, chronic fatigue, headaches, muscle, and joint pain or sleep disturbances. In terms of neuropsychiatric symptoms, 21 (27.3%) exhibited symptoms of anxiety, and 30 (39%) reported depressive symptoms. Regarding perceived stress, 15 patients (19.5%) reported rarely or never feeling stressed, 36 patients (46.7%) experienced stress occasionally, and 26 patients (33.8%) reported often or usually feeling stressed. Scores for PHQ-15 (**Fig. 1b**), PSS (**Fig. 1c**), HADS-A (**Fig. 1d**), and HADS-D (**Fig. 1e**) showed no significant differences across COVID-19 Severity Index groups, nor did they vary significantly with age or BMI (**Table 2**). Additionally, there were no significant differences in the prevalence of somatic, stress, or depressive symptoms between male and female patients. However, anxiety symptoms were more prevalent among women, who also scored higher on the HADS-A (**Table 2**). The prevalence of fever was higher among those with severe (PHQ ≥15) somatic symptoms (χ^2^(1)=4.586; p=0.032). There were significant correlations between PHQ-15 and PSS (**Fig. 1f**), HADS-A (**Fig. 1g**), and HADS-D (**Fig. 1h**). An association was found between stress, anxiety, and depressive symptoms scores. Higher stress levels were correlated with more extended hospital stays (r_s_=0.227; p=0.047).

**Fig. 1.**
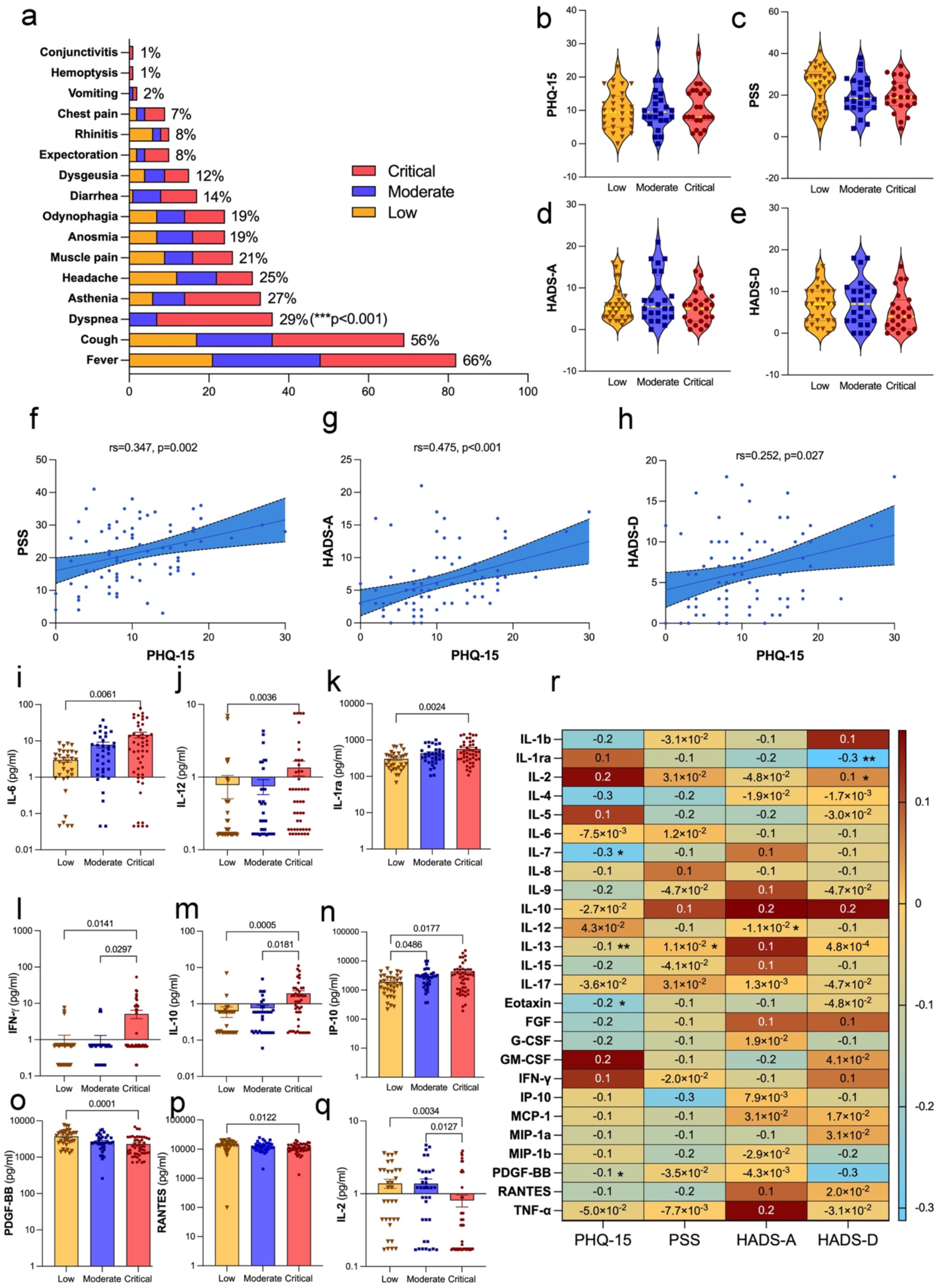
COVID-19 symptom prevalence, somatic and neuropsychiatric assessments in hhospitalized COVID-19 patients. (a) The prevalence of COVID-19 symptoms during hospitalization is categorized into low, moderate, and critical severity groups. The most common symptoms were fever (66%), cough (56%), and dyspnea (29%) which was more prevalent in the critical group (p<0.001). The chi-square or Fisher’s exact test was used to analyze contingency tables. Median and interquartile range of the clinical outcomes results of the patients classified according to the COVID-19 Severity Index (n=77). **(b)** Somatic symptoms (PHQ-15, *Patient Health Questionnaire-15*), **(c)** perception of stress *(PSS, Perceived Stress Scale),* **(d)** anxiety, and **(e)** depression (HADS-D, *Hospital Anxiety and Depression Scale)* scores were the same across the three groups categorized by COVID-19 severity (low, moderate, and critical). Pairwise comparisons between severity groups followed the Kruskal-Wallis test. Spearman’s Rank Correlation Coefficient showed a positive correlation between PHQ-15 scores and **(f)** PSS scores (r_s_=0.347, p=0.002), **(g)** HADS-A scores (r_s_=0.475, p<0.001) and **(h)** HADS-D scores (r_s_=0.252, p=0.027). These results indicate that higher somatic symptom burden is associated with increased perceived stress, anxiety, and depressive symptoms in hospitalized COVID-19 patients. The concentrations (pg/ml) of **(i)** IL-6, **(j)** IL-12, and **(k)** IL-1ra were higher in the critical group compared to the low group. For **(l)** IFN-γ and **(m)** IL-10, the critical group showed higher levels compared to the low and moderate groups. The levels of **(n)** IP-10 were higher in the critical and moderate groups compared to the low group. Conversely, the levels of **(o)** PDGF-BB, and **(p)** RANTES decreased from the low group to the critical group. For **(q)** IL-2, the critical group showed lower levels than the low and moderate groups. Pairwise comparisons between the severity groups followed the Kruskal-Wallis test. The lines and p-values on the Fig.s indicate significant differences between these groups. Linear regression was used to compare variables with and without adjustment for age, gender, and BMI. Age influenced IFN-γ serum concentration. Both age and BMI affected the differences in IL-12 and RANTES levels. For the rest of the cytokines (IL-1ra, IL-2, IL-6, IL-10, IP-10, PDGF-BB), the factor responsible for the differences in concentrations was the severity of COVID-19. **(r)** Heatmap illustrating the correlations between various cytokine levels in serum concentration (% coefficient of variation) and somatic and neuropsychiatric symptoms scores in hospitalized COVID-19 patients, with significant correlations marked with asterisks (*p<0.05, **p<0.01, ***p<0.001). Red shades represent positive correlations, whereas blue shades represent negative correlations. The Kruskal-Wallis test was followed by pairwise comparisons between severity groups with correction for multiple testing. IL, Interleukin; IL-1ra, Interleukin-1 receptor antagonist; IFN-γ, Interferon-gamma; IP-10, Interferon-gamma-inducible protein 10; PDGF-BB, Platelet-Derived Growth Factor BB; RANTES, Regulated on Activation, Normal T Cell Expressed and Secreted.

**Table 2.** Somatic and neuropsychiatric symptoms of the participants (n=77).

| Variables | PHQ-15 |  | PSS |  | HADS-A |  | HADS-D |  |
| --- | --- | --- | --- | --- | --- | --- | --- | --- |
|  | Median<br>(IQR) | *p*<br>*Value* | Median<br>(IQR) | *p*<br>*Value* | Median<br>(IQR) | *p*<br>*Value* | Median<br>(IQR) | *p*<br>*Value* |
| Gender |  |  |  |  |  |  |  |  |
| *Female* | 10 (7 – 14.5) | 0.393 | 23 (14 – 31.5) | 0.332 | 6 (3.5 – 13) | 0.014 | 7 (3 – 10.25) | 0.216 |
| *Male* | 8 (6.25 – 15) |  | 19 (15 – 27.5) |  | 4 (2 – 6.75) |  | 5 (1.25 – 9.75) |  |
| Age |  |  |  |  |  |  |  |  |
| *<60 years* | 9.5 (5.75 – 15) | 0.369 | 20.5 (14 – 28.25) | 0.398 | 5 (3 – 8) | 0.487 | 6 (3 – 10) | 0.751 |
| *≥60 years* | 8 (8 – 16) |  | 22 (16 – 31) |  | 6 (3 – 14) |  | 5 (2 – 12) |  |
| BMI |  |  |  |  |  |  |  |  |
| *<30 kg/m<sup>2</sup>* | 8 (5 – 14) | 0.222 | 22 (14 – 28) | 0.919 | 5 (3 – 7) | 0.536 | 6 (3 – 10) | 0.707 |
| *≥30 kg/m<sup>2</sup>* | 10 (7.75 – 16) |  | 19.5 (16 – 29.25) |  | 6 (3 – 9) |  | 5.5 (2 – 10) |  |
| COVID-19 severity |  |  |  |  |  |  |  |  |
| *Low* | 9.5 (5 – 14.25) | 0.864 | 27 (14.5 – 31.25) | 0.172 | 5 (3 – 8.25) | 0.496 | 7 (3 – 10) | 0.431 |
| *Moderate* | 9.5 (7 – 13.75) |  | 18.5 (14 – 26.75) |  | 5.5 (3 – 13) |  | 6 (2.5 – 10.5) |  |
| *Critical* | 8 (7 – 16) |  | 19 (15 – 26) |  | 5 (2 – 7) |  | 4 (1 – 8) |  |

**Note:** Comparison of scores on instruments assessing somatic symptoms, stress, anxiety, and depressive symptoms according to gender, age, BMI, and COVID-19 severity. Abbreviations: BMI, Body Mass Index; HADS-A, *Hospital Anxiety and Depression Scale-Anxiety*; HADS-D, *Hospital Anxiety and Depression Scale-Depression*; IQR, interquartile range; PHQ-15, *Patient Health Questionnaire-15*; PSS, *Perception of stress*. Mann-Whitney U test or Kruskal-Wallis test was followed by pairwise comparisons between groups.

The levels of the inflammatory mediators – interleukin (IL) (IL-6, IL-12, IL-10), IL-1 receptor antagonist (IL-1ra), interferon-gamma (IFN-γ), and interferon-gamma-inducible protein 10 (IP-10) were higher with increased severity of SARS-CoV-2 infection (**Fig. 1i-n**). On the other hand, platelet-derived growth factor BB (PDGF-BB), regulated on activation, normal T cell expressed and secreted (RANTES), and interleukin-2 (IL-2) were lower in critically ill patients (**Fig. 1o-q**). There were no differences in the other measured inflammatory mediators (**Supplementary** Fig. 1). The scores obtained in the PHQ-15 were associated with interleukin-7 (IL-7), interleukin-13 (IL-13), eotaxin, and PDGF-BB levels. PSS and IL-13 were correlated. Anxiety scores were correlated with IL-12. The scores obtained in the HADS-D were associated with IL-1ra and IL-2. The heatmap displays the statistically significant correlations (**Fig. 1r**).

Additionally, the number of neutrophils in circulation increased in moderate and critical patients compared to those with low COVID-19 severity, whereas circulating lymphocytes decreased. An increase in the NLR and PLR accompanied these changes. CRP, D-dimer, fibrinogen, ferritin, and LDH were elevated with COVID-19 severity. Albumin concentration decreased in moderately severe and critically ill patients (**Supplementary** Fig. 2).

### Impact of COVID-19 severity on gut microbiome composition: associations with BMI, gender, and age

The analysis of alpha diversity (Shannon index) (**Fig. 2a**) revealed a significant reduction in gut microbiota diversity in critically ill COVID-19 patients compared to those with low disease severity (p<0.05). Regarding species richness (Chao1 index) (**Fig. 2b**), critically ill patients showed significantly lower richness compared to those with low (p<0.01) and moderate (p<0.05) disease severity. Beta diversity analyses (MDS plots) demonstrated that BMI significantly correlated the gut microbiota composition in patients classified as having low COVID-19 severity (**Fig. 2c-e**), with a distinct separation between patients with BMI ≥30 and those with BMI under 30. Gender-based analysis (**Fig. 2f-h**) significantly correlated microbiota composition only in critically ill patients (**Fig. 2h**). Similarly, age was significantly associated with microbiota composition in the critically ill group, with patients ≥60 years showing distinct microbial profiles compared to those under 60 years (**Fig. 2k**). Bar plots of microbial relative abundance at the family (**Fig. 2l**) and genus levels (**Fig. 2m**) demonstrated considerable shifts in microbial composition across severity groups.

**Fig. 2.**
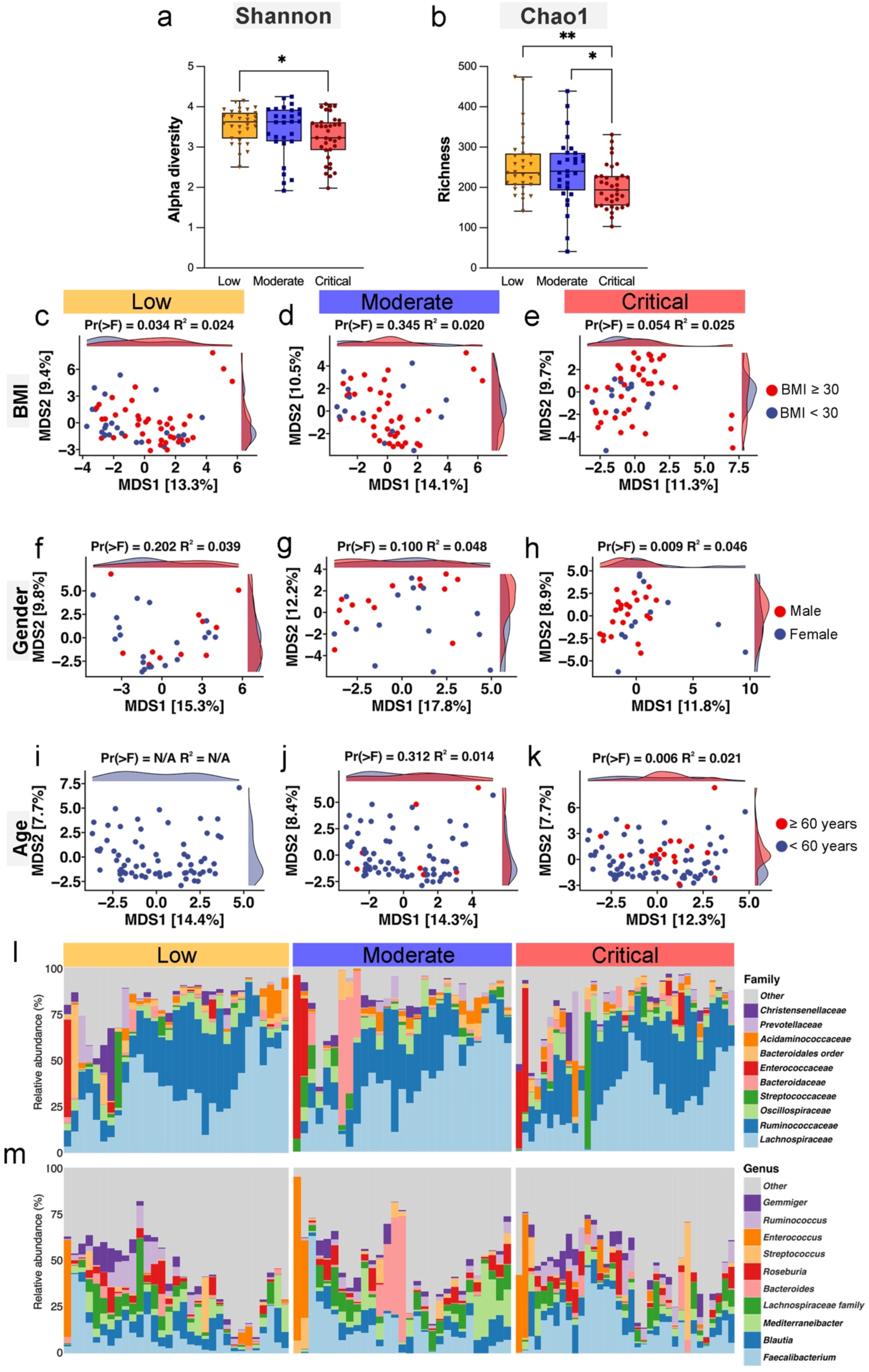
Alpha and beta diversity of gut microbiome in hospitalized COVID-19 patients. **(a)** Shannon index, which measures species richness and evenness, indicating lower diversity in the critical group compared to the low group (p=0.0129) (**b**) Chao1 richness estimator indicates significantly lower species counts in the critical group compared to the low group (p=0.0073) and the moderate group (p=0.0197) (*p<0.05 and **p<0.01). (**c-e**) Beta diversity analysis was performed using Aitchison Principal Coordinates Analysis (PCoA) for Body-mass-index (BMI) categories (<30 vs. ≥30 kg/m^2^), showing distinct clustering in the low group (p=0.034). (**f-h**) PCoA plots for gender (female vs. male) show distinct clustering in the critical severity group (p=0.009), suggesting gender-based differences in microbiome composition are more pronounced in this group. (**i-k**) PCoA plots for age categories (<60 vs. ≥60 years) show a more distinct clustering in the critical group (p=0.006), indicating age-related differences in microbiome composition. The statistical significance of these groupings was assessed using permutational multivariate analysis of variance (Permanova) within the vegan R package, with Pr(>F) and R^2^ values confirming the observed differences in microbial community composition. The relative abundance of various gut microbiota showing the top 15 taxa at family level (**l**) and genus level (**m**) in hospitalized COVID-19 patients across different severity groups.

### Differential gut microbial signatures across COVID-19 severity

Analysis of the Composition of Microbiomes with Bias Correction 2 (ANCOMBC2) can potentially uncover biomarkers or therapeutic targets by providing a list of bacteria with significant changes in abundances. The heatmap (**Fig. 3a**) shows the centered log-ratio abundance of gut microbial species across patients with low, moderate, and critical COVID-19 severity. A clear shift in microbial composition was observed as disease severity increased, with certain species becoming more abundant in critically ill patients while others decreased. The graphs within **Fig. 3b** and **Fig. 3c** show log fold changes of specific bacterial species between low to moderate and low to critical severity groups, respectively. Notably, species such as *Streptococcus periodonticum* and *Clostridium perfringens* were significantly enriched in patients with lower disease severity (q<0.001 and q<0.01) (**Fig. 3b** and **Fig. 3c)**. In comparison, *Klebsiella pneumonia* and *Prevotella loescheii* were highly abundant in critically ill patients (q<0.001 and q<0.01) (**Fig. 3c**).

**Fig. 3.**
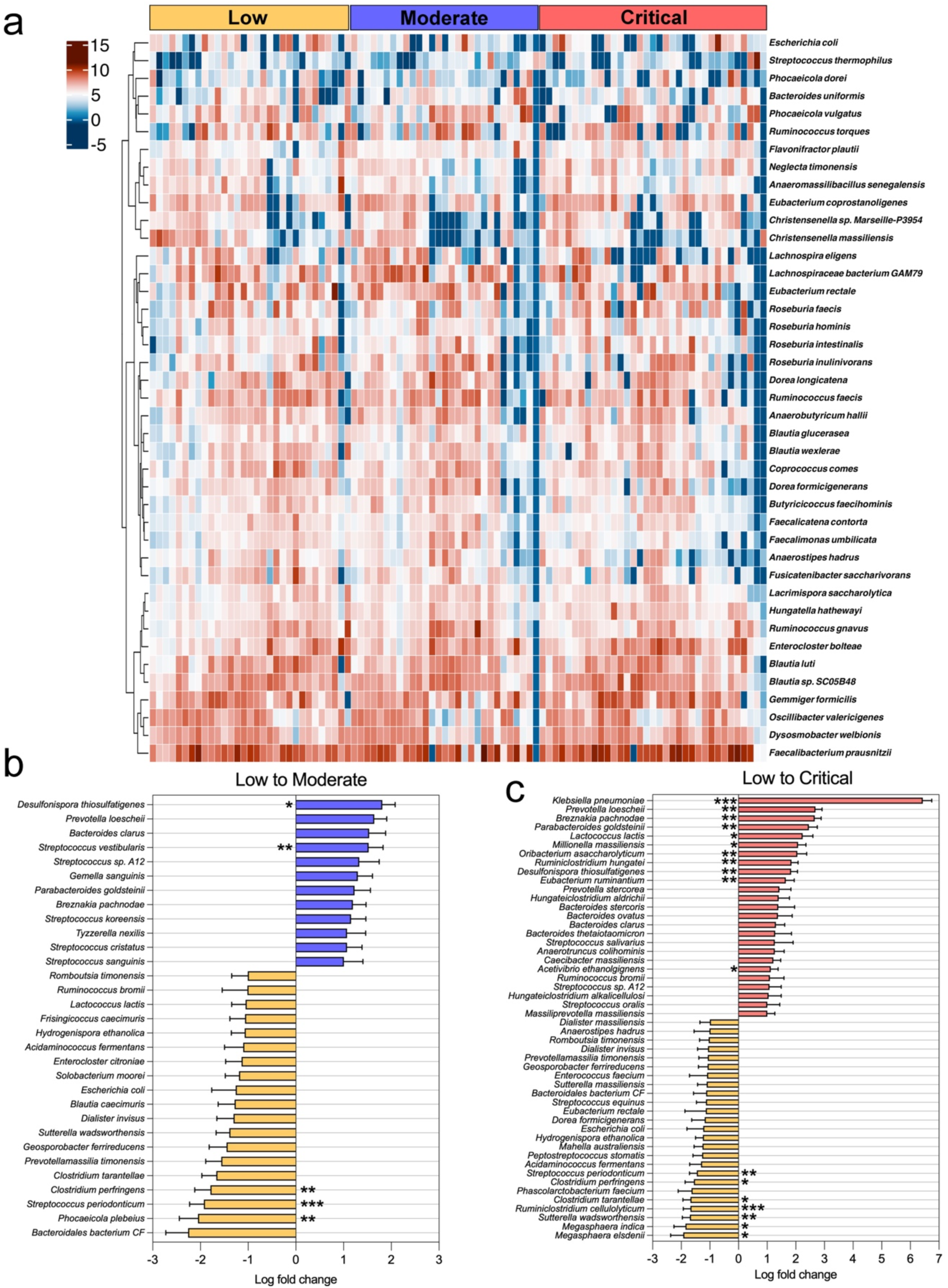
Heatmap and differential abundance of specific gut microbiota taxa in hospitalized COVID-19 patients across different severity groups. **(a)** Heatmap of the centered-log transformed abundance of significantly altered taxa showing intra-group variation among patients with the same COVID-19 severity, highlighting distinct microbial profiles associated with each group. The color intensity represents the transformed abundance, with darker shades of red indicating higher abundance. **(b)** Comparison between low and moderate-severity groups reveals significant increases in taxa such as *Desulfonispora thiosulfatigenes* and *Streptococcus vestibularis* in moderate group. It decreases in taxa such as *Streptococcus periodonticum, Phocaeicola plebius,* and *Clostridium perfringens* in moderate group. **(c)** Comparison between low and critical severity groups shows significant increases in taxa such as *Klebsiella pneumoniae, Prevotella loeschii*, and *Breznakia pachnodae* with corresponding decreases in *Ruminiclostridium cellulolyticum, Streptococcus periodonticum,* and *Suterella wadsworthensis* in critical group; identified using ANCOMBC2. The analysis revealed differentially abundant species with statistical significance levels indicated as *q<0.05, **q<0.01, and ***q<0.001.

### Association of gut microbiota with somatic symptoms, stress, anxiety, and depressive symptoms in COVID-19 patients

Principal Coordinates Analysis (PCoA) was performed to investigate associations between somatic and neuropsychiatric symptoms and gut microbiome composition in patients with COVID-19 (**Fig. 4a-l**). A statistically significant association was found between somatization, as measured by the PHQ-15, and microbiome composition within the moderate severity group (p=0.04, R²=0.029; **Fig. 4b**). This finding suggests that individuals with higher somatic symptom scores may exhibit distinct microbial profiles compared to those with lower scores. Similarly, perceived stress showed a non-significant trend with the microbiome in the moderate group (p=0.05, R²=0.084; **Fig. 4e**), indicating that experiencing stress regularly could influence microbial community composition. The relatively higher R² value for PSS suggested that perceived stress explained a notable portion of the observed variance in microbial composition compared to other mental health factors. In contrast, scores from the HADS-A and HADS-D tests did not display significant associations with microbiome composition across all severity groups (p>0.05; **Figs. 4g–5l**). These results suggest that, in this sample, general measures of anxiety and depressive symptoms may not significantly contribute to variations in gut microbial structure.

**Fig. 4.**
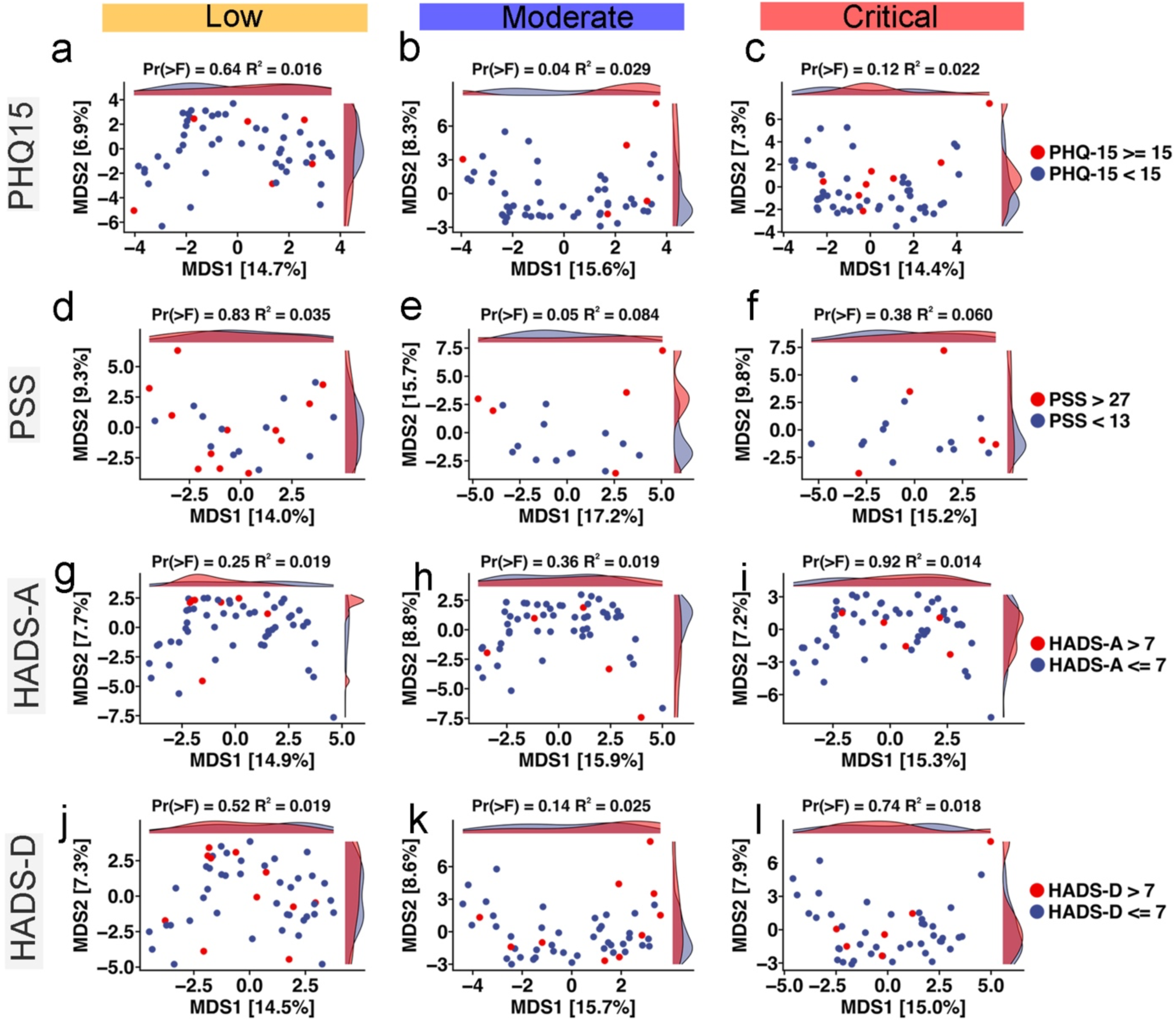
Ordination analyses of gut microbiota composition, somatic and neuropsychiatric symptoms across COVID-19 severity levels. **(a-c)** Aitchison distance principal coordinate analysis plots explore the relationships between COVID-19 severity, gut microbiota composition, somatic symptoms (PHQ-15) **(d-f)**, and neuropsychiatric symptoms, including stress (PSS) **(a-l)**, anxiety (HADS-A) **(g-i)**, depressive symptoms (HADS-D) **(j-l)**. The distributions indicate varying degrees of correlation between gut microbiota diversity and mental health indicators, with Permanova (>F) and R² values specified for each plot.

**Fig. 5.**
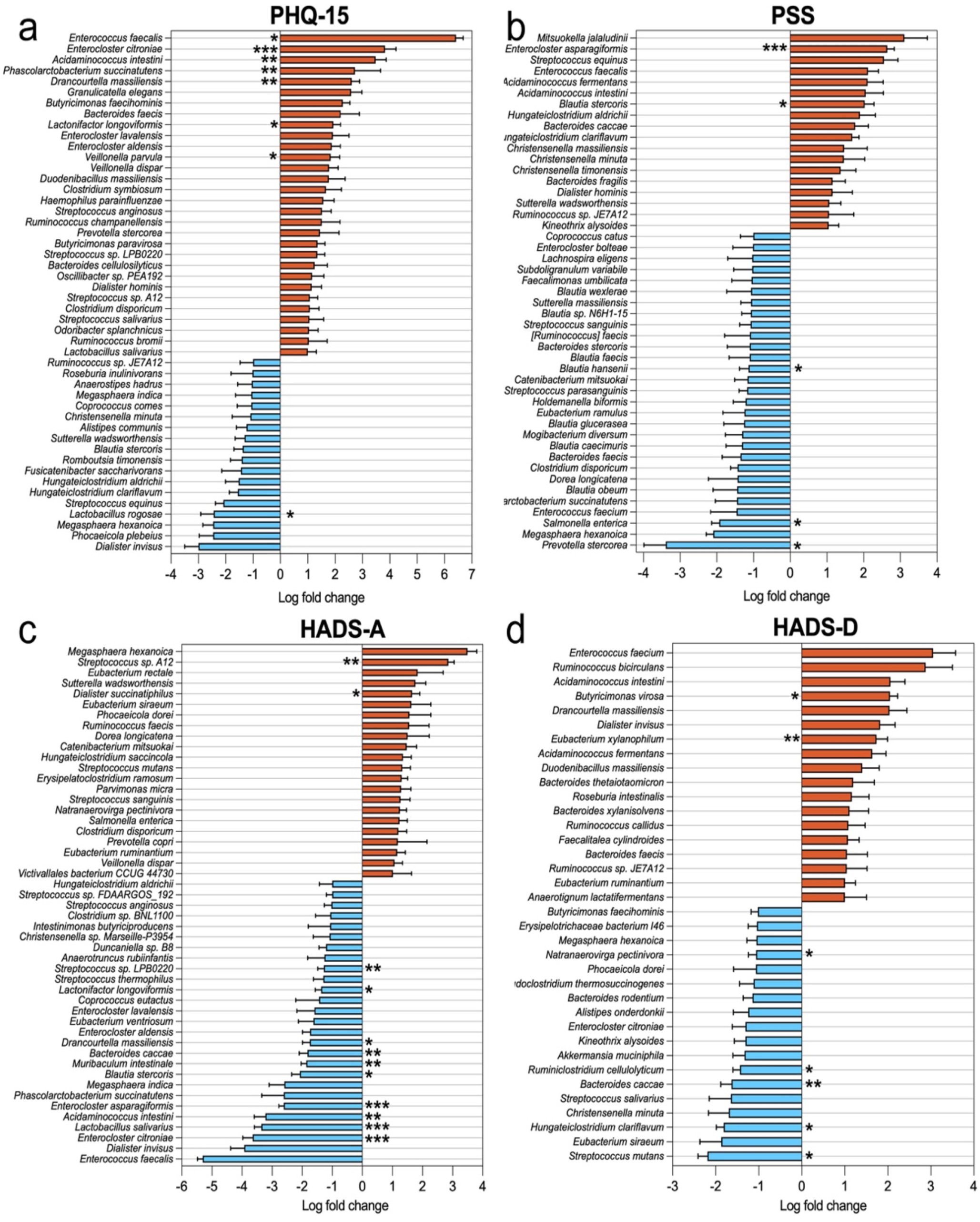
Differential abundance analysis between gut microbiota, somatic and neuropsychiatric symptoms in hospitalized COVID-19 patients. Log fold changes in abundance of various bacterial taxa associated with four different assessments: **(a)** PHQ-15 (somatic symptoms), **(b)** PSS (perceived stress), **(c)** HADS-A (anxiety), and **(d)** HADS-D (depressive symptoms). The analysis used ANCOMBC2 to identify differentially abundant microbes between high-and low-test scores. Bars represent log fold changes in microbial abundance, with red bars indicating an increase and blue bars indicating a decrease in abundance associated with higher test scores. Error bars represent standard errors calculated using ANCOMBC2. Asterisks (*q<0.05, **q<0.01, and ***q<0.001) denote statistical significance levels, with more asterisks indicating higher significance. Only the top differentially abundant taxa are shown for each scale (LFC ζ1).

A secondary differential abundance analysis revealed microbial shifts correlated with scores on the PHQ-15, PSS, HADS-A, and HADS-D assessments. Patients with higher PHQ-15 scores showed a significant increase in the abundance of species such as *Enterococcus citroniae* (q<0.001), *Phascolarctobacterium succinatutens and Acidaminococcus intestine* (q<0.01), and *Enterococcus faecalis* (q<0.05), while known beneficial bacteria like *Lactobacillus rugosae* were significantly depleted (q<0.05) (**Fig. 5a**). Higher PSS scores were associated with an overrepresentation of *Enterocloster asparagiformis* (q<0.001), and *Blautia stercoris* (q<0.05). At the same time, species *Blautia hansenii*, *Salmonella enterica*, and *Prevotella stercorea* (q<0.05) were significantly decreased (**Fig. 5b**). HADS-A scores were linked to an increase in *Dialister succinatiphilus (q<0.05) and Streptococcus sp. A12 (q<0.01)*, among others, showed a notable reduction in species such as *Lactobacillus salivarius* (q<0.001) and *Bacteroides caccae* (q<0.01) (**Fig. 5c**). Higher HADS-D were correlated with elevated levels of *Butyricimonas virosa* (q<0.05), *and Eubacterium xylanophilum* (q<0.01). In contrast, species *Bacteroides caccae* (q<0.01) and *Ruminiclostridium cellulolyticum* (q<0.05) were significantly diminished (**Fig. 5d**).

### Distinct gut microbial correlations with somatic symptoms, stress, anxiety, and depressive symptoms in COVID-19 patients

The heatmap presents the Spearman correlation coefficients of CLR-transformed abundances between various bacterial species and four assessments: PHQ-15, PSS, HADS-A, and HADS-D (**Fig. 6**). Several microbial species showed significant correlations with these symptoms, with only associations where p<0.05 displayed numerically to highlight statistically significant relationships. For instance, *Christensenella minuta* (p<0.01), *Anaeromassilibacillus senegalensis, Lachnospiraceae bacterium GAM79*, and *Christensenella massiliensis* (p<0.05) were negatively associated with somatic symptoms (PHQ-15), showing a significant reduction. Conversely, *Mitsuokella jalaludinii, Prevotella copri,* and *Phocaeicola plebeius* were positively associated with increased PHQ-15 scores (p<0.05). Notably, *Mitsuokella jalaludinii* showed significant positive correlations with all assessments (p<0.05)), indicating a potential association between its increased abundance and higher symptom burden. Additionally, *Phocaeicola vulgatus* correlates with PSS and HADS-D (p<0.05). *Bacteroides stercoris* correlates with HADS-D (p<0.05).

**Fig. 6.**
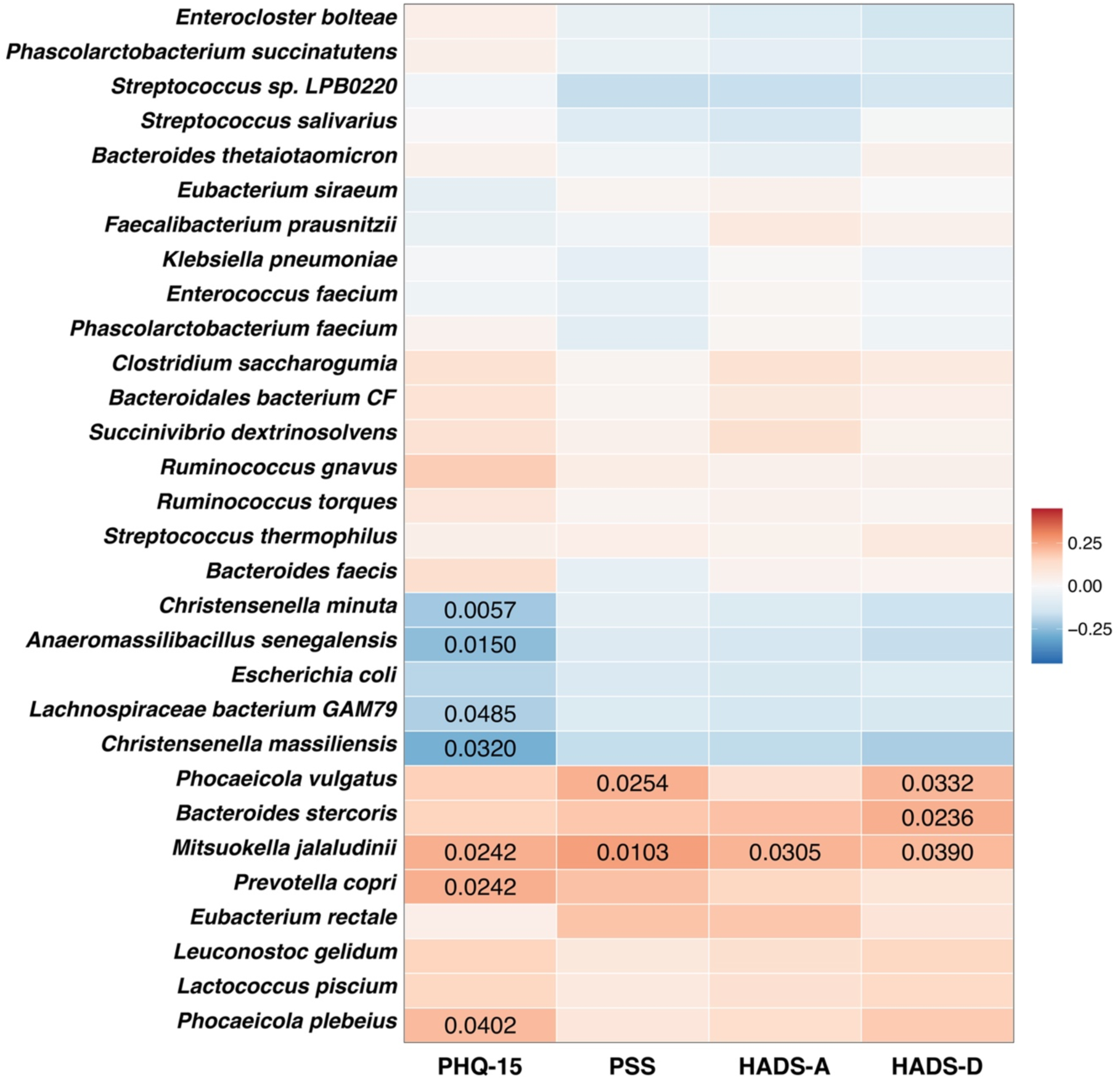
Heatmap of correlations between gut microbiota species, somatic and neuropsychiatric symptoms. Heatmap illustrating the Spearman’s correlation coefficients between the centered log-ratio (CLR) transformed abundances of gut microbial species and scores from four assessments: PHQ-15 (somatic symptoms), PSS (perceived stress), HADS-A (anxiety), and HADS-D (depression). The color scale represents the strength and direction of correlations, with red indicating positive correlations and blue indicating negative correlations. The intensity of the color corresponds to the magnitude of the correlation, as shown in the legend on the right (ranging from -0.25 to 0.25). P-values are less than 0.05 displayed on corresponding cells, indicating statistical significance. Blank cells represent correlations that did not meet the significance threshold. Bacterial species are listed on the y-axis, while the mental health scales are shown on the x-axis. This visualization allows for identifying specific microbial species that may have consistent or unique associations with different aspects of mental health as measured by these indicators. Notable correlations include positive associations between *Mitsuokella jalaludinii* and all measurements, and negative associations between several species (e.g., *Christensenella massiliensis, Lachnospiraceae bacterium GAM79*) and PHQ-15 scores.

### Multivariate analysis and network visualization of gut microbial associations with somatic and neuropsychiatric symptoms in COVID-19 patients

To further explore the relationship among somatic and neuropsychiatric symptoms, COVID-19 severity, and gut microbiome composition, a redundancy analysis (RDA) was conducted. This analysis visualized the distribution of microbial species with COVID-19 severity (low, moderate, critical) and all assessments (PHQ-15, PSS, HADS-A, HADS-D). The RDA plot (**Fig. 7a**) shows the relationship between gut microbial species and PHQ-15, PSS, HADS-A, and HADS-D scores across different COVID-19 severity groups. The RDA axes account for 4.9% and 1.4% of the variation, respectively. Notably, the species *Mitsuokella jalaludinii* was positioned within the mental health indicator constraints of the plot, suggesting potential associations with specific assessed symptoms or COVID-19 severity outcomes. Distinct relationships were observed, with species such as *Phocaeicola vulgatus*, *Bacteroides stercoris,* and *Prevotella copri* being positively associated with elevated somatic scores, particularly in patients with critical disease.

**Fig. 7.**
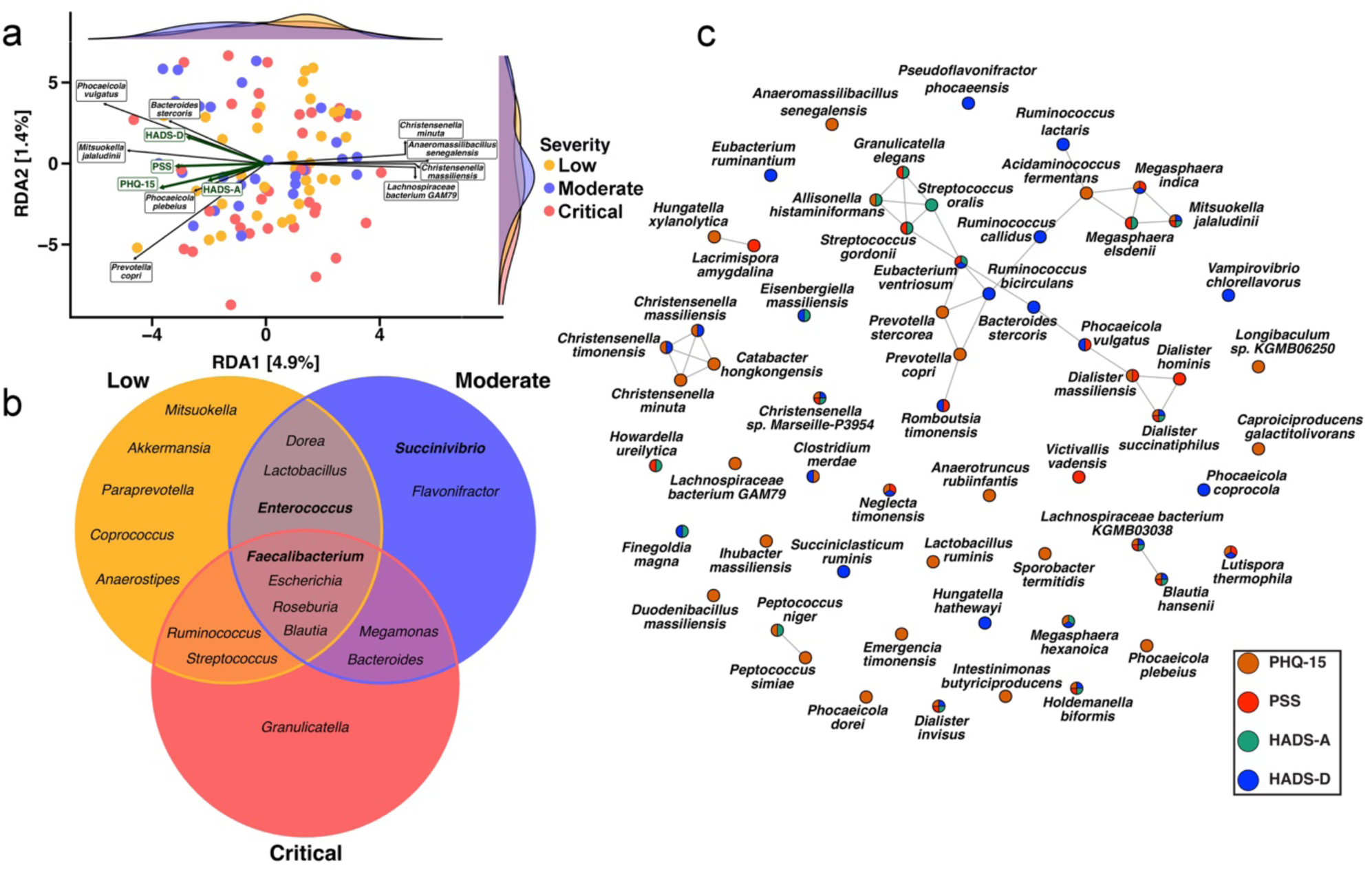
Associations between gut microbiota, somatic and neuropsychiatric symptoms in hospitalized COVID-19 patients. **(a)** Redundancy Analysis (RDA) plot showing the association between gut microbiome composition, somatic and neuropsychiatric symptoms, and COVID-19 severity. The x-axis represents RDA1 (4.9% of variation), and the y-axis represents RDA2 (1.4%). Each point represents an individual patient, color-coded by COVID-19 severity. Green arrows indicate the direction and strength of association with PHQ-15, PSS, HADS-A, and HADS-D. Labeled bacterial species show their associations with these factors. Density plots on the top and right margins show the distribution of samples along each axis. **(b)** Venn diagram of the resulting driver taxa from each COVID-19 severity group using the BakDrive interaction analysis followed by the BakDrive driver analysis. Genera that are bolded correspond to taxa that are above the total relative abundance threshold of 5%. **(c)** SpiecEasi, sparse and low-rank decomposition network analysis of gut microbiome species, illustrates the co-occurrence patterns of gut bacterial species associated with mental health indicators. Nodes represent bacterial species with a p<0.05 with any mental health index, while edges represent significant correlations between species. Node colors represent Spearman’s correlation between the microbe and the scores of the assessments. Together, these visualizations provide insights into the complex relationships between gut microbiome composition, mental health status, and COVID-19 severity, suggesting potential microbial signatures associated with different symptoms in the context of COVID-19 infection.

Conversely, species *Christensenella minuta, Anaeromassilibacillus senegalensis,* and *Lachnospiraceae bacterium GAM79* negatively correlated with somatic symptoms, suggesting a potential protective effect. To investigate which microbial taxa might drive the severity of COVID-19 in gut microbiomes, we used Bakdrive(*25*). Bakdrive is a microbial community modeling tool designed to identify driver bacteria that play a pivotal role in influencing the structure and function of the microbiome in health or disease. Specifically, by utilizing metabolic models and network analysis, Bakdrive aims to pinpoint taxa that play crucial roles in disease mechanisms or ecological interactions. Using this tool, we can highlight the bacterial species that were likely driving interactions within gut microbiomes across low, moderate, and critical severity groups. Specific taxa exhibited varying prevalence across different levels of COVID-19 severity (**Supplementary Table 3**). For example, *Granulicatella* was predominantly present in critically ill patients, while *Succinivibrio* was more common among those with moderate disease severity. Some driver taxa were shared between severity groups. Notably, *Faecalibacterium* was present across all severity levels, and *Enterococcus* appeared only in patients with low and moderate severity. These findings reflect distinct variations in driver taxa composition associated with disease severity scores (**Fig. 7b**).

The network diagram generated using SpiecEasi (Sparse InversE Covariance estimation for Ecological Association Inference) illustrated the associations between specific microbial species and various symptoms assessed, highlighting Spearman correlations with p-values <0.05 related to mental health indicators (**Fig. 7c**). This network highlights the intricate interactions among bacterial species, with nodes color-coded to reflect their associations with PHQ-15, PSS, HADS-A, and HADS-D scores. The structure reveals positive and negative associations between species, computed using the sparse and low-rank decomposition method. This approach minimizes the impact of latent variables, allowing for a more precise representation of the complex balance within the gut microbiome with mental health and COVID-19 severity. Microbial species such as *Mitsuokella jalaludinii* and *Phocaeicola vulgatus* strongly correlated with PSS and HADS-D. Conversely, species *Christensenella minuta, Anaeromassilibacillus senegalensis, and Lachnospiraceae bacterium GAM79* were inversely associated with PHQ-15. Key species *Ruminococcus lactaris, Megasphaera indica*, and *Bacteroides stercoris* occupy central positions within the network, indicating their potential significance in the gut-brain axis and their possible role in influencing mental health status.

## Discussion

This proof-of-concept study highlights links between somatic and neuropsychiatric symptoms, inflammatory responses, and gut microbiome dysbiosis in hospitalized COVID-19 patients. The results provide essential insights into the intricate interactions between COVID-19 severity and specific microbial species, demonstrating that different microbes are associated with varying levels of disease severity and neuropsychiatric co-morbidities.

In this study, most patients experienced at least three COVID-19 symptoms, with dyspnea significantly associated with increased disease severity. Consistent with previous research, older age, and male gender were linked to more severe manifestations of COVID-19, reaffirming age as a significant risk factor for severe outcomes(*4, 5*). Somatic symptoms, stress, anxiety, and depressive symptoms were common across all severity levels. This aligns with other studies reporting a high prevalence of physical symptoms and mental health issues among hospitalized COVID-19 patients during the same period(*10, 11, 26, 27*). Notably, a higher proportion of women reported anxiety and had higher scores on the HADS-A scale compared to men. Individuals with higher stress levels also experienced more extended hospital stays. These findings are consistent with studies identifying women as a vulnerable group for mental health issues, including perceived stress(*26*) and anxiety(*27*).

Our results align with existing literature regarding alterations in blood cells and markers among critically ill patients, such as reduced lymphocytes, elevated neutrophils, increased CRP, D-dimer, and ferritin levels, and comorbidities like hypertension and heart diseases (*28*). These findings underscore the significant role of systemic inflammation and pro-thrombotic responses in exacerbating COVID-19 severity(*29–31*). SARS-CoV-2 infection triggers immunopathological reactions characterized by intense cytokine production, known as the cytokine storm(*32*). Elevated levels of IL-6, as observed in our study, are frequently reported, especially in individuals over 60 years old with comorbidities, and are recognized as markers of disease severity storm(*32*). Elevated IL-6 is associated with increased production of acute-phase proteins like CRP and other inflammatory cytokines(*29–31*). In line with other findings, IL-1ra, IL-10, and IFN-γ were significantly elevated in patients with severe COVID-19(*30*). The increase in IL-10 and IL-1ra may reflect a compensatory anti-inflammatory response to elevated pro-inflammatory cytokines, attempting to mitigate the harmful effects of the cytokine storm(*33*). However, this response appears ineffective, as elevated IL-10 levels are associated with poor clinical outcomes and reduced survival rates(*33, 34*). Additionally, we observed decreased levels of IL-2 in critically ill patients, which may represent a response aimed at preventing viral spread during the early phase of the disease(*35*).

An inverse relationship was observed between disease severity and the abundance of beneficial bacteria. Alpha diversity and species richness were significantly reduced in critically ill patients, reinforcing previous findings of microbiome dysbiosis in severe COVID-19 cases(*36*). This diversity reduction aligns with findings linking diminished gut microbial diversity to poor symptoms and heightened inflammatory responses(*37*). Consistent with this, beta diversity analyses revealed distinct microbial profiles influenced by factors such as BMI, gender, and age, as reported by other studies (*38*), highlighting the impact of these demographic factors on the gut microbiome during COVID-19 infection. Severely ill patients exhibited a decline in beneficial bacterial taxa, including *Christensenella minuta* and *Lachnospiraceae,* which are known to support gut health, and their reduction suggests a weakening of the intestine’s protective functions. This could lead to dysbiosis, a decrease in short-chain fatty acids (SCFA), crucial for maintaining intestinal integrity and regulating immune responses, ultimately contributing to poorer mental health outcomes in severely ill patients. Another study during the same period found that severely symptomatic SARS-CoV-2 patients had significantly lower bacterial diversity and reduced abundances of beneficial bacteria, such as *Bifidobacterium*, *Faecalibacterium*, and *Roseburium* while showing an increase in *Bacteroides* compared to controls(*39*). Several bacterial taxa associated with SARS-CoV-2 infection were identified, notably elevated levels of *Granulicatella* and *Rothia mucilaginosa* found in both the oral and gut microbiomes(*40*). Consistent with our findings, *Granulicatella* was exclusively predominant in critical patients.

An increase in opportunistic pathogens, like *Klebsiella pneumoniae* and *Parabacteroides distasonis*, can intensify inflammation, raise infection risks, and potentially worsen a patient’s overall health (*41*). For instance, *Klebsiella pneumoniae* was commonly associated with critically ill patients and linked to severe conditions like pneumonia and sepsis(*41*). Certain pathogens have also been associated with specific symptoms in long-term COVID-19; for example*, Clostridium innocuum* has been suggested to play a role in neuropsychiatric symptoms(*42*). Many gut microbiome taxa associated with known comorbidities, such as *Veillonella dispar*, *Veillonella parvula*, and *Streptococcus gordonii*, are typically found within the oral cavity. Their presence in the gut aligns with several other studies showing a link between disease severity and oral taxa within the gut microbiome(*43*). Bacterial genera such as *Eubacterium, Agathobacter, Subdoligranulum, Ruminococcus, and Veillonella* show notable shifts in abundance.

Interestingly, we observed that specific microbial species were associated with somatic symptoms, perceived stress, anxiety, and depressive symptoms, offering insights into the complex microbial dynamics, the gut-brain axis, and mental health indicators in COVID-19. The gut microbiome influences neuropsychiatric symptoms through microbial metabolites like SCFA, which regulate immune cells and neurotransmitter production, and through the gut-brain axis(*44*). Imbalances in gut microbes can lead to increased inflammation and disrupted communication, contributing to conditions like anxiety, depressive symptoms, and cognitive decline(*44, 45*). Furthermore, the gut microbiome has been suggested to be a key modulator of psychological health during and after COVID-19 infection (*37, 45*).

Differential abundance analyses showed that several microbial species were significantly associated with psychological measures, including somatic symptoms, perceived stress, anxiety, and depressive symptoms. The clustering of species like *Phocaeicola vulgatus, Bacteroides stercoris, Ruminococcus lactaris, Megasphaera indica*, and *Mitsuokella jalaludinii* around elevated somatic or neuropsychiatric scores suggests that these microbes may play a vital role in the gut-brain axis during COVID-19. *Mitsuokella jalaludinii* was positively correlated with all assessments, suggesting its role in worsening mental health(*46*). In contrast, species like *Christensenella minuta* and *Anaeromassilibacillus senegalensis* were negatively associated with PHQ-15. These findings align with growing evidence that links gut dysbiosis to mood disorders and stress-related conditions.

Limitations of this study include its cross-sectional design, which prevents establishing causality between gut microbiome alterations and neuropsychiatric symptoms. Potential confounding factors such as pre-existing mental health conditions or medication use were not controlled for, and a lack of a healthy control group limits the ability to compare findings to non-COVID-19 individuals. Additionally, a larger sample size may be required to generalize the results to all hospitalized COVID-19 patients. Finally, fecal microbial load may represent a major confounder (*47*).

Recovery from SARS-CoV-2 infection has been linked to greater psychosocial resilience (*27*), while individuals with higher levels of mental distress during the acute phase are more vulnerable to long-term effects (*48*). Hospitalized patients who experienced severe COVID-19 face a greater risk of developing Long COVID compared to those with mild cases(*49*). Our meta-analysis, published in 2021, found that 8 out of 10 individuals with SARS-CoV-2 infections exhibited symptoms of Long COVID(*50*). However, recent epidemiological data offer a more conservative estimate, Long COVID risk at three months post-infection is now reported at 6.2%(*49, 51*). It is very important to consider parameters with strong discriminatory power to stratify patient risk and classify the severity of COVID-19(*52*). Randomized controlled trials are necessary to assess the effectiveness of microbiome-targeted therapies, such as probiotics and dietary interventions. Mechanistic studies should investigate how microbiome changes influence systemic inflammation during the acute phase of COVID-19 and how these interactions may predict the onset and progression of neuropsychiatric symptoms. This research may help guide the development of clinical guidelines and patient monitoring strategies for incorporating microbiome-based interventions into COVID-19 treatment.

In conclusion, our study provides data in support of the hypothesis that immune responses and the gut-brain axis may be playing a role in regulating somatic and neuropsychiatric symptoms during acute COVID-19. Probiotics, prebiotics, and dietary modifications, such as increasing fiber or omega-3 intake to restore microbial balance, could reduce the severity of symptoms. Additionally, microbiome-targeted therapies or pharmaceutical approaches that modulate gut health offer promising avenues to improve recovery and mitigate the risk of severe complications in COVID-19 patients. Further research is needed to explore whether modulating the gut microbiota could alleviate neurological symptoms during the infection phase and impact the development of Long COVID.

## Materials and Methods

### Study Design

This cross-sectional study was based on an exploratory analysis of baseline data from hospitalized COVID-19 patients who participated in a randomized clinical trial investigating the effects of a dietary supplement containing tannins(*53*). The study protocol has been detailed in a prior report(*54*). The study was approved by the Ethics Committee of the Hospital de Clínicas José de San Martín (1046–20) in accordance with the guidelines of the Argentine Ministry of Health and the hospital’s treatment protocols. The demographics and clinical characteristics of the participants are presented in **Table 1**, and the methodology is outlined in **Fig. 9**. This study was performed between 1 March 2020 and 31 October 2021.

**Fig. 9.**
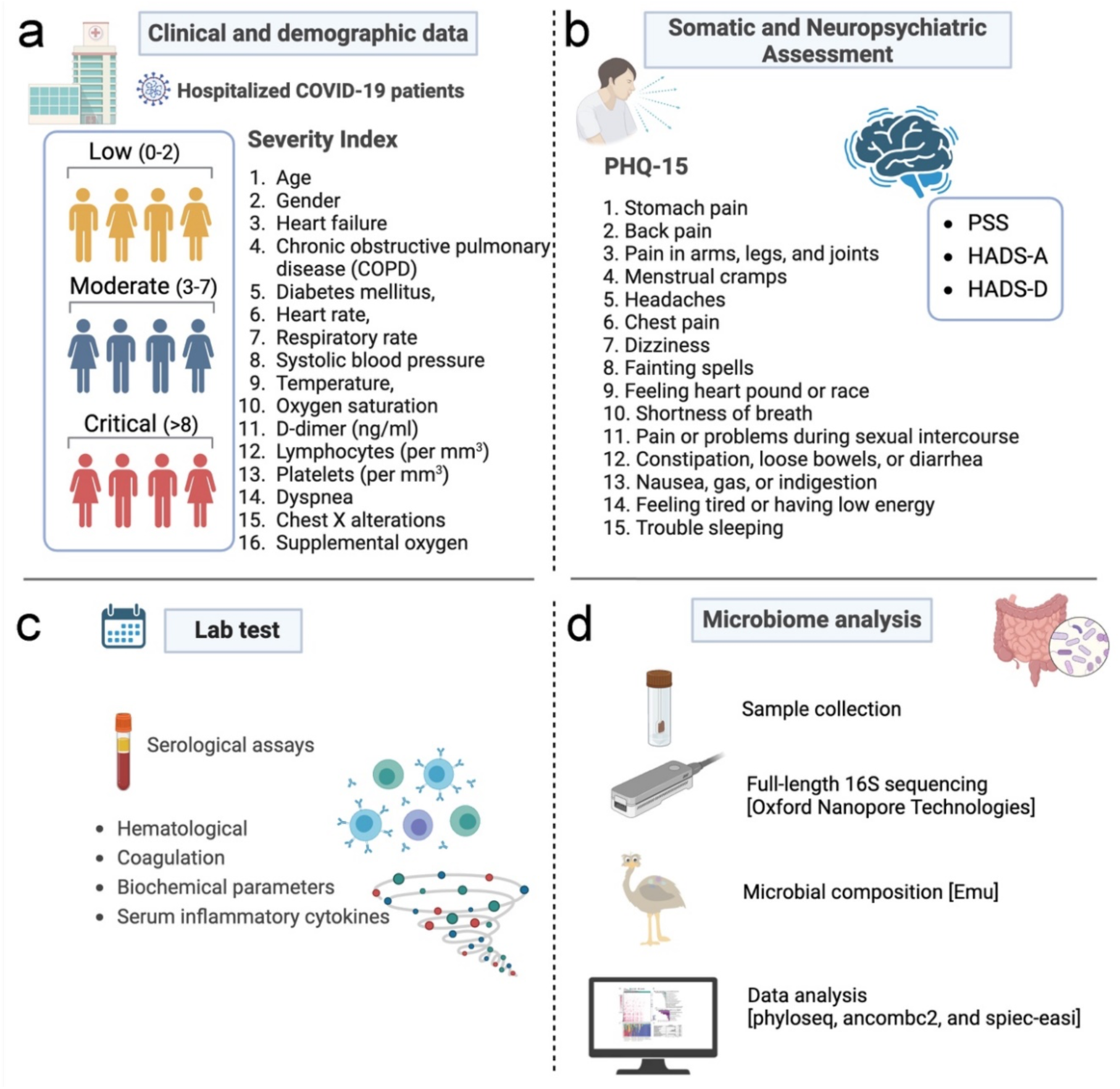
Overview of data collection and analysis for hospitalized COVID-19 patients. This comprehensive data collection framework aims to provide a holistic understanding of the COVID-19 symptoms and somatic, neuropsychiatric, and microbiological aspects of COVID-19 in hospitalized patients. (a) Clinical and Demographic Data. COVID patients (n=124) are categorized into three severity groups based on a scoring index: low (0–2), moderate (3–7), and critical (>8). This score is based on age, gender, heart failure, chronic obstructive pulmonary disease (COPD), diabetes mellitus, heart rate, respiratory rate, systolic blood pressure, temperature, oxygen saturation, including in patients with COPD, D-dimer levels, lymphocyte and platelet counts, dyspnea, chest X-ray alterations, and the need for supplemental oxygen. (b) Somatic and Neuropsychiatric Assessments: Patient Health Questionnaire-15 (PHQ-15) is used to assess somatic symptoms, including stomach pain, back pain, pain in arms, legs, and joints, menstrual cramps, headaches, chest pain, dizziness, fainting spells, heart palpitations, shortness of breath, pain during sexual intercourse, constipation, nausea, indigestion, fatigue, and trouble sleeping. Additional neuropsychiatric assessments include the Perceived Stress Scale (PSS), Hospital Anxiety and Depression Scale-Anxiety (HADS-A), and Hospital Anxiety and Depression Scale-Depression (HADS-D). (c) Laboratory Tests. Various serological assays are conducted to analyze hematological, coagulation, and biochemical parameters, and serum inflammatory cytokines. (d) Microbiome Analysis. Stool samples are collected for microbiome analysis. Full-length 16S sequencing is performed using Oxford Nanopore Technologies, followed by microbial composition analysis using Emu. Data analysis uses tools such as phyloseq, ANCOMBC2, and SpiecEasi to understand the microbial landscape and its association with disease severity and patient outcomes.

### COVID-19 severity classification

Using the COVID-19 severity index, patients were classified into low, moderate, or critical disease severity(*55*). This index incorporates the following multiple variables: age, gender, comorbidities, dyspnea, chest X-ray abnormalities, and the requirement for supplemental oxygen (**Fig.9**). The X-ray was performed at the time of hospital admission. Here, we considered scores of 0–2 to indicate low severity, 3–7 as moderate, and 8 or higher as critical severity of COVID-19 adapted from Huespe et al. (*56*) (**Supplementary Table 1**).

### Somatic and neuropsychiatric symptoms

Somatic symptoms were assessed using the Patient Health Questionnaire-15 (PHQ-15), while the perception of stress was evaluated with the Perceived Stress Scale (PSS). Anxiety and depressive symptoms were measured using the Hospital Anxiety and Depression Scale (HADS), with all somatic and neuropsychiatric symptoms being self-reported. Only 77 patients could complete the instruments, which was influenced by the severity of the clinical condition and transfer to the intensive care unit at the time of hospital admission. The PHQ-15 consists of 15 items that assess the severity of somatic symptoms over the previous week(*57*) (**Fig. 9**). Each symptom is scored as 0 (“not bothered at all”), 1 (“bothered a little”), or 2 (“bothered a lot”). The PHQ-15 score ranges from 0 to 30, (0-28 in men) with cutoff points at ≥5 for low, ≥10 for moderate, and ≥15 for high somatic symptom severity(*57*). Scores of 15 or higher are indicative of severe somatic symptoms. The PSS(*58*) is a self-report measure that assesses the perception of stress over the previous month. The version of the PSS used was the 14-item version, which includes seven positive and seven negative items that assess feelings of chaos, lack of control, and overall stress without being tied to specific events. Each item is rated on a 5-point scale from 0 (“never”) to 4 (“very often”), with higher scores indicating more significant stress. The categories are rarely or never (less than 14 points); occasionally (14-28 points); often (29-42 points); and usually (42-56 points). The HADS(*59*) is a self-report rating scale of 14 items designed to measure anxiety and depression (7 items for each subscale). Each item is scored from 0 (“absence”) to 3 (“extreme presence”). Each subscale has a total score ranging from 0 to 21, with scores of 0–7 indicating “normal,” 8–10 “mild,” 11–14 “moderate,” and 15–21 “severe” symptoms.

### Blood biomarkers

Blood samples were collected by venipuncture within 48 hours of hospital admission and placed in vacutainer tubes with a clot activator. Serum was obtained by centrifugation at 3,000× g for 15 minutes at 4°C. Circulating cytokine levels were evaluated using the Bio-Plex Pro™ Human Cytokine Standard 27-Plex Kit (Bio-Rad), following the manufacturer’s instructions. This assay measures 27 different molecules: IL-2, IL-4, IL-5, IL-6, IL-7, IL-8, IL-9, IL-10, IL-12, IL-13, IL-15, IL-17, IL-1β, IL-1ra, TNF-α, IFN-γ, IP-10, MCP-1, MIP-1α, MIP-1β, eotaxin, FGF, G-CSF, GM-CSF, VEGF, and PDGF-BB. Serum levels of the inflammatory mediators were reported as median fluorescence intensity (MFI). Counts of leukocytes, neutrophils, lymphocytes, and platelets were measured. The neutrophil-to-lymphocyte ratio (NLR) and platelet-to-lymphocyte ratio (PLR) were subsequently calculated. Coagulation parameters assessed included prothrombin time, activated partial thromboplastin time (aPTT), D-dimer, and fibrinogen levels. Inflammatory markers, specifically C-reactive protein (CRP) and ferritin were evaluated to analyze inflammatory status. Additionally, biochemical analyses were conducted to determine the concentrations of albumin, creatine kinase (CK), and lactate dehydrogenase (LDH).

### Nanopore 16S rRNA library preparation and sequencing

Stool samples were collected using μb-eNAT preservation tubes (COPAN®, Italy), aliquoted, and stored at −80 °C until analysis. DNA was extracted from 100 mg of homogenized fecal samples employing the MagMAX™ Microbiome Ultra Nucleic Acid Isolation Kit, following the manufacturer’s protocol. Subsequently, 16S rRNA amplicon libraries were prepared using the 16S Barcoding Kit 1–24 (SQK-16S024) from Oxford Nanopore Technologies (ONT), Oxford, UK. For PCR amplification and barcoding, 18 ng of extracted DNA was combined with LongAmp Hot Start Taq 2X Master Mix (New England Biolabs, Ipswich, MA) according to the manufacturer’s instructions. The thermal cycling protocol consisted of an initial denaturation at 95 °C for 20 seconds, followed by 35 cycles of 95 °C for 20 seconds, 55 °C for 30 seconds, and 65 °C for 2 minutes, concluding with a final extension at 65 °C for 5 minutes. The resulting barcoded amplicons were purified using AMPure XP beads (Beckman Coulter, Brea, CA). After purification, the amplicons were quantified with a Qubit fluorometer (Life Technologies, Carlsbad, CA) and pooled in an equimolar ratio to achieve 100 ng in 10 µL. The pooled library was then loaded into an R9.4.1 flow cell and sequenced on the MinION platform (Oxford Nanopore Technologies, Oxford, UK) using MINKNOW software version 19.12.5 for data acquisition.

### Bioinformatic Analysis

Full-length 16S sequences were converted to pod5 format and called using Dorado v0.5.0 with the dna_r9.4.1_e8_sup@v3.6 model. The resulting BAM files were demultiplexed and converted to fastq files using the Dorado demux command(*60*). Fastq files for each sample were then concatenated using SeqKit scat v2.7.0, processed with the Emu taxonomic taxonomic classification software v3.4.5, and aggregated into an Operational Taxonomic Unit (OTU) output table(*61*). The OTU and taxonomy tables were integrated with metadata to create a phyloseq v1.48.0 object(*62*). A phylogenetic tree was created using the Emu taxdump_to_tree.py script to generate a reference tree based on the Emu database. Using a custom Python script (https://github.com/villapollab/covid_biome) and a mapping file specific to the dataset, leaves that matched within the reference tree were retained and converted to their corresponding OTU identifiers in the phyloseq object. The phylogenetic tree was merged with the phyloseq object using the ape v5.8 read.tree function, enabling phylogenetic calculations within phyloseq(*63*). Subsequent analyses included alpha diversity, beta diversity, and abundance and correlation heatmaps, performed using MicroViz v0.12.4(*64*). Network correlations were obtained using SpiecEasi version 1.1.3(*65*). All data analyses following Emu classification were executed using Python v3.12 and R v4.3.3(*66, 67*).

### Differential Abundance and Driver Species Analysis

To obtain a comprehensive understanding of potential bacterial driver species within the gut microbial communities of COVID-19 patients of differing severity, we employed Bakdrive v1.0.4(*25*) and ANCOMBC2 v2.6.0(*68*). Bakdrive is a microbial community modeling tool designed to identify driver bacteria—bacteria that play a pivotal role in influencing the structure and function of the microbiome in health or disease. Specifically, by utilizing metabolic models and network analysis, Bakdrive attempts to pinpoint taxa that may be critical in disease mechanisms or ecological interactions. ANCOMBC2 (Analysis of Composition of Microbiomes with Bias Correction 2) is a differential abundance analysis tool designed to address the compositional nature of microbiome data and corrects for biases, enabling accurate identification of taxa that are differentially abundant between sample groups. A detailed notebook of the bioinformatic analysis can be found at https://villapollab.github.io/covid_biome/.

### Statistical Analysis

Categorical variables were presented as frequencies and percentages. To analyze contingency tables for variables such as gender, mortality, antibiotic use, supplemental oxygen, and COVID-19 symptoms, the Chi-square (χ²) test or Fisher’s exact test (FET) was employed. The normality of the distribution of numerical variables was assessed with a Kolmogorov-Smirnov test. Depending on their distribution, numerical variables were presented as median and interquartile range (IQR) or mean and standard deviation (SD). For comparisons between two groups, the Independent Samples t-test was used for normally distributed variables, while the Mann-Whitney U test was applied for non-normally distributed variables (e.g., PSS, HADS-A, HADS-D between groups with and without severe somatic symptoms). One-way ANOVA was utilized to examine differences in parametric variables such as age and BMI across COVID-19 severity categories (low, moderate, and critical), followed by Tukey’s Honest Significant Difference (HSD) post hoc test. For non-parametric variables, including days of hospitalization, number of COVID-19 symptoms, PHQ-15, PSS, HADS-A, and HADS-D, the Kruskal-Wallis test was performed. This was followed by pairwise comparisons between severity groups with correction for multiple testing. Linear regression analysis compared cytokine levels with and without adjustment for age, gender, and BMI. Associations between clinical outcomes and inflammatory mediators or gut microbiota were assessed using Spearman’s rank correlation coefficients (e.g., correlations between PHQ-15, PSS, HADS-A, and HADS-D). All statistical analyses were performed using IBM SPSS Statistics for Windows, version 25 (IBM Corp., Armonk, NY, USA). A p-value of <0.05 was considered statistically significant.

## Data Availability

All data are available in the main text or the supplementary materials. The raw sequences generated in this study have been deposited in the NCBI Sequence Read Archive (SRA) database under accession PRJNA1179392.

## Acknowledgments

The authors would like to thank Silvia Molino, Andrea Pisarevsky, Fabiana López Mingorance, Patricia Vega, Julieta Repetti, Guillermina Ludueña, Pablo Pepa, and Tatiana Uehara for their collaboration in the data and sample collection at the Hospital de Clínicas José de San Martín, Buenos Aires, Argentina. This work also received funding from Indunor/Silvateam SA. PS is supported by a fellowship from the Coordenação de Aperfeiçoamento de Pessoal de Nível Superior, Brazil (CAPES, Finance Code 001). A.M. is supported by a training fellowship from the Gulf Coast Consortia on the NLM Training Program in Biomedical Informatics & Data Science (T15LM007093). The Houston Methodist NeuralCODR Fellowship program supports SS. Fig. 9a was created using BioRender.

## Author contributions

SV, PS, AM, SS, KC, TJT, MMP, and SV prepared the manuscript. SV supervised the study. MD, TH, and EMQ commented critically on the manuscript and data interpretation. MMP contributed to participant recruitment and sample collection. AM, SS, KC, and TJT contributed to the computational analysis of the de-identified microbiome sequencing data. PS and SV contributed to the study design, data interpretation, analysis, and manuscript writing. All authors gave final approval for the version to be published.

## Data and materials availability

All data are available in the main text or the supplementary materials. The raw sequences generated in this study have been deposited in the NCBI Sequence Read Archive (SRA) database under accession PRJNA1179392. All bioinformatic data are available on Github: https://github.com/villapollab/covid_biome/releases/tag/medrxiv_sub.

