## Supplemental Material for "Gut microbiome dysbiosis and immune activation correlate with somatic and neuropsychiatric symptoms in COVID-19 patients"

Paula Scalzo *et al.*

**This PDF file includes:**

Figs. S1 to S2
Tables S1 to S3

### Supporting figures

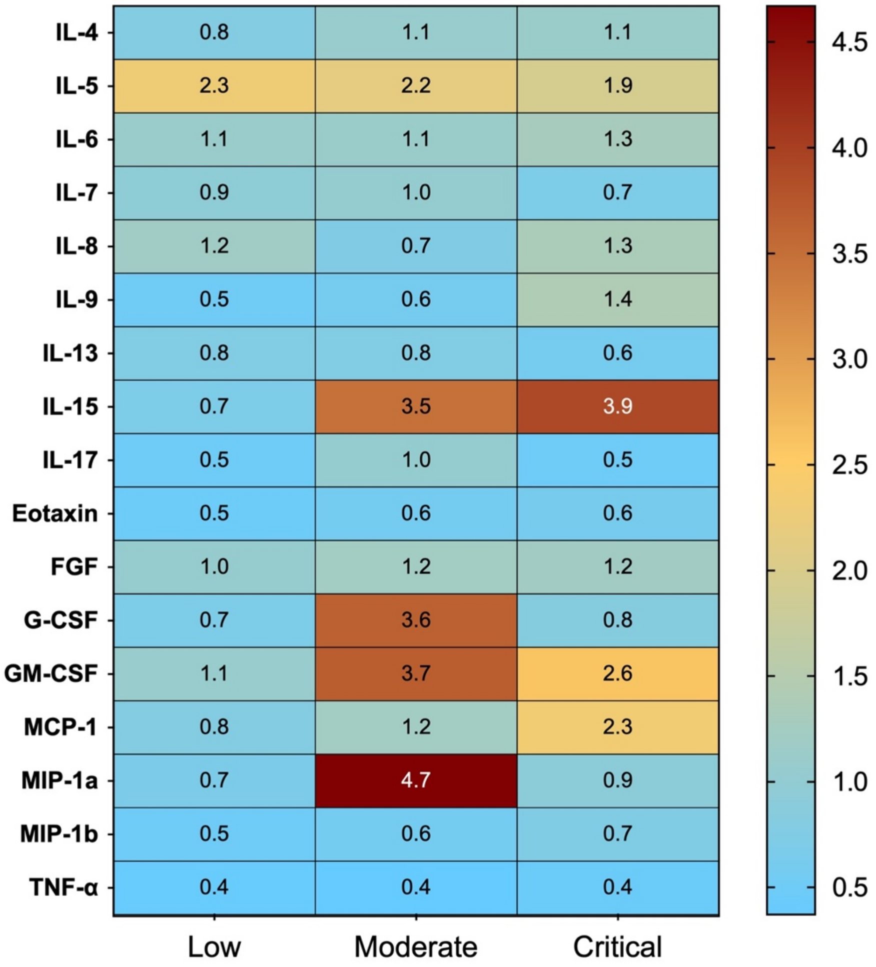


**Fig. S1. Cytokine and Chemokine Profiles Across COVID-19 Severity Levels.** Cytokine and chemokine levels across COVID-19 severity groups. Heatmap depicting the relative concentrations of cytokines and chemokines in patients with low, moderate, and critical COVID-19 severity. Significant differences between severity groups are marked (*p < 0.05, **p < 0.01, ***p < 0.001). Elevated levels of IL-1ra, IL-6, IL-10, IL-12, IL-15, and MIP-1a are observed in moderate and critical cases, highlighting their role in systemic inflammation and disease progression.

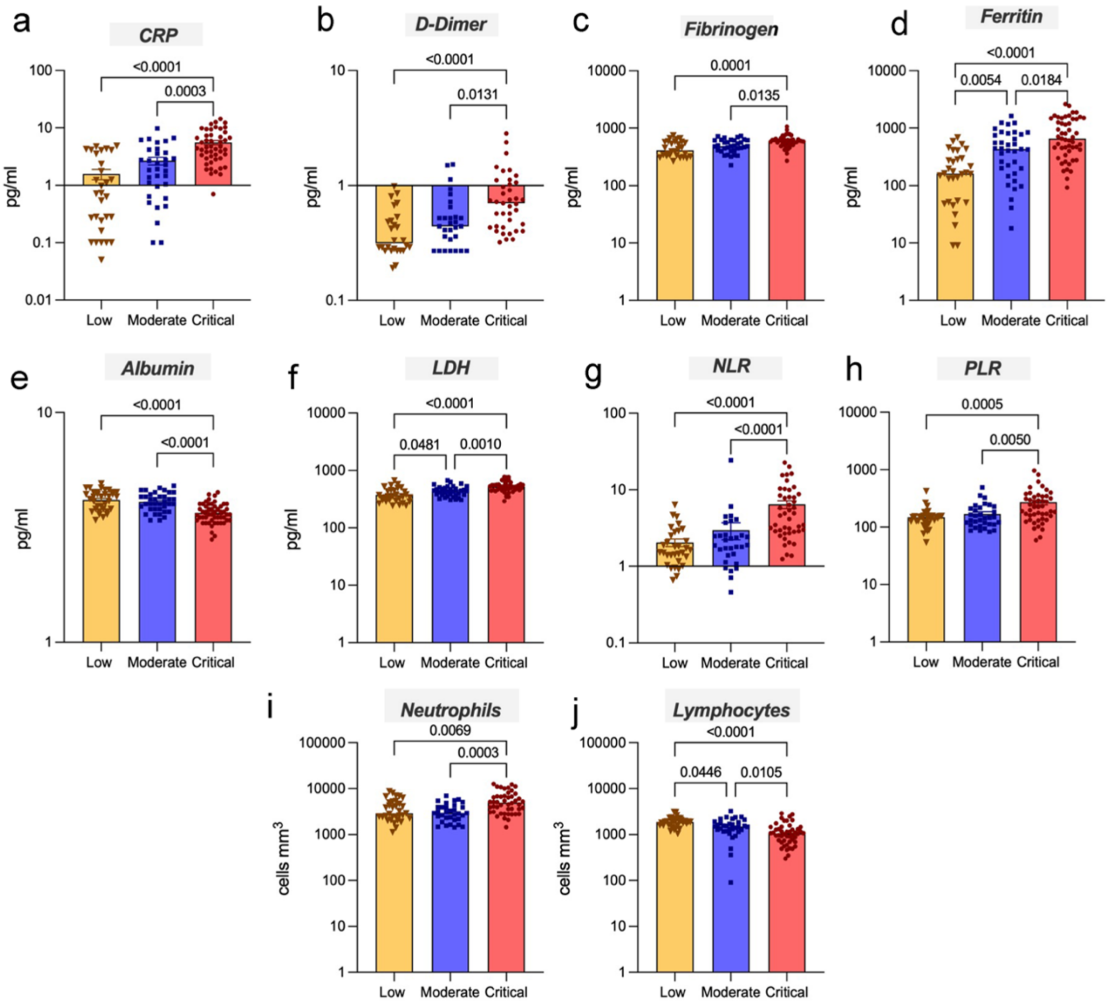


**Fig. S2. Alterations in inflammatory and hematological markers in hospitalized COVID-19 patients across different severity groups.** The levels of (a) CRP, (b) D-dimer, (c) Fibrinogen, as well as (d) Neutrophil counts, (e) Neutrophil-to-lymphocyte ratio (NLR), (f) Platelet-to-lymphocyte ratio (PLR) were elevated in patients from the critical group compared to those in the low and moderate groups. Besides the elevated levels in the critical group compared to the low and moderate groups, the levels in the moderate group were also higher compared to the low group for the concentrations of (g) Ferritin, and (h) Lactate dehydrogenase (LDH). Conversely, (i) Albumin levels decrease with increasing severity, with significant reductions in the critical group compared to low and moderate groups. (j) Lymphocyte counts decrease with increasing severity, showing lower counts in critical patients compared to patients in low and moderate groups. These values were lower in moderate compared to low group. ANOVA was followed by Šidak multiple comparisons test or Kruskal-Wallis test was followed by pairwise comparisons between the severity groups. The lines and p-values on the Fig.s indicate significant differences between these groups.

### Supporting tables

**Table S1**. COVID-19 Index Severity.

| **Variables** | | **Scores** |
|---|---|---|
| **Demographic data** | Age (years) | ≤60 = 0<br>61-64 = 1<br>≥65 = 2 |
| | Gender | Female = 0 / Male = 1 |
| **Chronic diseases** | Heart failure | No = 0 / Yes = 1 |
| | COPD | No = 0 / Yes = 1 |
| | Diabetes mellitus | No = 0 / Yes = 1 |
| **Vital signs** | Heart rate (beats per minute) | ≤40 = 3<br>41 – 50 = 1<br>51 – 90 = 0<br>91 – 110 = 1<br>111 – 130 = 2<br>≥131 = 3 |
| | Respiratory rate (breaths per minute) | ≤8 = 3<br>9 – 11 = 1<br>12 – 20 = 0<br>21 – 24 = 2<br>≥25 = 3 |
| | Systolic blood pressure (mmHg) | ≤90 = 3<br>90 – 219 = 0<br>≥220 = 3 |
| | Temperature (°C) | ≤35 = 3<br>35.1 – 35.5 = 1<br>35.6 – 37.9 = 0<br>38 – 39 = 1<br>≥39.1 = 2 |
| | Oxygen saturation (%) | ≤91 = 3<br>92 – 93 = 2<br>94 – 95 = 1<br>≥96 = 0 |
| | Oxygen saturation in patients with COPD (%) | ≤83 = 3<br>84 – 85 = 2<br>86 – 87 = 1<br>≥88 = 0 |
| **Laboratory tests** | D-dimer (ng/ml) | ≤1000 = 0<br>>1000 = 1 |
| | Lymphocytes (per $mm^3$) | ≥1000 = 0<br><1000 = 1 |
| | Platelets (per $mm^3$) | ≥10000 = 0<br><10000 = 1 |
| **Imaging test** | Chest X-ray | Normal = 0<br>Bilateral infiltration = 1 |
| **Clinical conditions** | Dyspnea | No = 0 / Yes = 2 |
| | Supplement oxygen | No = 0 / Yes = 3 |

Predictive variables of worse outcome at hospital admission. Scores of 0-2 indicates low severity, 3-4 moderate, 5-7 high, and 8 or above indicates critical COVID-19 (Adapted from Huespe et al., 2020). COPD: Chronic obstructive pulmonary disease.

**Table S2.** Hematological, coagulation, inflammatory, and biochemical parameters of the patients classified according to the COVID-19 Severity Index (n=124).

| Variables | All patients (n=124) | COVID-19 Severity | | | |
|---|---|---|---|---|---|
| | | Low (n=34) | Moderate (n=37) | Critical (n=53) | p Value |
| Hematological parameters | | | | | |
| *Leukocytes (cells/µL)* | 5660 (4690 – 8120) | 5640 (4750 – 8205) | 5045 (4505 – 7078) | 6430 (4765 – 9410) | 0.085 |
| *Neutrophils (cells/µL)* | 3825 (2513 – 5438) | 2880 (2330 – 5040) | 2930 (2135 – 3955) | 4955 (3115 – 6808) | 0.0001 |
| *Neutrophils / Lymphocytes ratio (NLR)* | 2.5 (1.5 – 4.6) | 1.6 (1.2 – 2.7) | 1.9 (1.4 – 2.6) | 4.6 (2.8 – 8.6) | <0.0001 |
| *Lymphocytes (cells/µL)* | 1460 (1040 – 1900) | 1880 (1590 – 2135) | 1495 (1178 – 1858) | 1055 (710 – 1423) | <0.001 |
| *Platelets (nº/µL)* | 242927 ± 79063 | 268647 ± 82137 | 230892 ± 66402 | 234673 ± 82824 | 0.080 |
| *Platelets / Lymphocytes ratio (PLR)* | 157 (121 – 245) | 147 (116 – 157) | 145 (110 – 220) | 224 (140 – 342) | 0.0002 |
| Coagulation parameters | | | | | |
| *Prothrombin time (s)* | 90.1 ± 9.9 | 89.3 ± 9.3 | 91.1 ± 10.2 | 89.9 ± 10.4 | 0.762 |
| *Activated partial thromboplastin time (s)* | 41.7 ± 6.1 | 41.8 ± 5.9 | 41.7 ± 5.9 | 41.7 ± 6.3 | 0.995 |
| *D-Dimer (µg/mL)* | 0.48 (0.34 – 0.76) | 0.32 (0.27 – 0.50) | 0.44 (0.31 – 0.59) | 0.70 (0.43 – 1.06) | <0.0001 |
| *Fibrinogen (mg/L)* | 548 (418 – 625) | 412 (330 – 576) | 484 (406 – 616) | 599 (516 – 643) | 0.0001 |
| Inflammatory parameters | | | | | |
| *C-reactive protein (mg/L)* | 2.93 (1.05 – 4.85) | 0.72 (0.22 – 3.67) | 1.92 (0.80 – 3.95) | 4.49 (2.94 – 7.45) | <0.0001 |
| *Ferritin (ng/mL)* | 433 (176 – 811) | 162 (51 – 296) | 433 (181 – 796) | 659 (399 – 1448) | <0.0001 |
| Biochemical parameters | | | | | |

| | | | | | |
|---|---|---|---|---|---|
| *Albumin (g/dL)* | 3.93 ± 0.44 | 4.17 ± 0.40 | 4.08 ± 0.38 | 3.65 ± 0.36 | <0.0001 |
| *Creatine kinase (UI/L)* | 94 (56 – 152) | 76 (54 – 135) | 98 (60 – 169) | 107 (48 – 156) | 0.372 |
| *Lactate dehydrogenase (UI/L)* | 456.4 ± 119.7 | 376.4 ± 107.7 | 438.5 ± 92.5 | 523.5 ± 109.6 | <0.0001 |

Note: The data were presented as median (interquartile range) or mean and standard deviation. ANOVA was followed by Šídak multiple comparisons test for parametric parameters or Kruskal-Wallis test was followed by pairwise comparisons between groups.

**Table S3.** The presence (1) and absence (0) of driver species between COVID-19 severity levels were identified using BakDrive.

| Bacteria Taxa | Low | Moderate | Critical |
|---|---|---|---|
| *Akkermansia muciniphila* | 1 | 0 | 0 |
| *Blautia obeum* | 1 | 1 | 0 |
| *Dorea formicigenerans* | 1 | 1 | 0 |
| *Enterococcus faecium* | 1 | 1 | 0 |
| *Escherichia coli* | 1 | 1 | 1 |
| *Faecalibacterium prausnitzii* | 1 | 1 | 1 |
| *Lactobacillus ruminis* | 1 | 0 | 0 |
| *Paraprevotella clara* | 1 | 0 | 0 |
| *Roseburia inulinivorans* | 1 | 1 | 1 |
| *Ruminococcus bromii* | 1 | 0 | 1 |
| *Streptococcus salivarius* | 1 | 0 | 0 |
| *Anaerostipes hadrus* | 1 | 0 | 0 |
| *Coprococcus catus* | 1 | 0 | 0 |
| *Coprococcus eutactus* | 1 | 0 | 0 |
| *Mitsuokella jalaludinii* | 1 | 0 | 0 |
| *Bacteroides faecis* | 0 | 1 | 0 |
| *Flavonifractor plautii* | 0 | 1 | 0 |
| *Lactobacillus mucosae* | 0 | 1 | 0 |
| *Megamonas funiformis* | 0 | 1 | 0 |
| *Succinivibrio dextrinosolvens* | 0 | 1 | 0 |
| *Bacteroides cellulosilyticus* | 0 | 0 | 1 |
| *Blautia producta* | 0 | 0 | 1 |
| *Blautia wexlerae* | 0 | 0 | 1 |

| **Bacteria Taxa** | **Low** | **Moderate** | **Critical** |
|---|---|---|---|
| *Granulicatella elegans* | 0 | 0 | 1 |
| *Megamonas hypermegale* | 0 | 0 | 1 |
| *Ruminococcus bicirculans* | 0 | 0 | 1 |
| *Ruminococcus champanellensis* | 0 | 0 | 1 |
| *Streptococcus thermophilus* | 0 | 0 | 1 |
| *Streptococcus sanguinis* | 0 | 0 | 1 |
